# Deep Learning for Pneumonia Diagnosis: A Custom CNN Approach with Superior Performance on Chest Radiographs

**DOI:** 10.1101/2025.05.26.25328342

**Authors:** Malay Vyas, Apurva A. Mehta

**Affiliations:** Dharmsinh Desai University, Computer Engineering, Nadiad 387001, India

**Keywords:** Convolutional Neural Network, Transfer Learning, Pneumonia detection, Chest X-ray images

## Abstract

A major global health and wellness issue causing major health problems and death, pneumonia underlines the need of quickly and precisely identifying and treating it. Though imaging technology has advanced, radiologists’ manual reading of chest X-rays still constitutes the basic method for pneumonia detection, which causes delays in both treatment and medical diagnosis. This study proposes a pneumonia detection method to automate the process using deep learning techniques. The concept employs a bespoke convolutional neural network (CNN) trained on different pneumonia-positive and pneumonia-negative cases from several healthcare providers. Various pre-processing steps were done on the chest radiographs to increase integrity and efficiency before teaching the design. Based on the comparison study with VGG19, ResNet50, InceptionV3, DenseNet201, and MobileNetV3, our bespoke CNN model was discovered to be the most efficient in balancing accuracy, recall, and parameter complexity. It shows 96.5% accuracy and 96.6% F1 score. This study contributes to the expansion of an automated, paired with a reliable, pneumonia finding system, which could improve personal outcomes and increase healthcare efficiency. The full project is available at here.

## 1 Introduction

Pneumonia ranks among the most prevalent etiologies of mortality in pediatric populations globally. In India, nearly 20% of these occurrences culminate in fatality [1]. Despite significant strides made internationally in mitigating the incidence of childhood pneumonia, it remains a substantial public health challenge [2,3,4]. A principal strategy to diminish the mortality rate involves ensuring timely diagnosis for affected individuals. Pneumonia possesses the potential to inflict damage upon one or both pulmonary structures. This damage may ensue as a consequence of infections instigated by bacteria, viruses, or fungi [5,6].

Pneumonia is characterized as an infection of the pulmonary system that elicits inflammation within lung cells and typically impacts the respiratory apparatus. Inhalation of harmful microorganisms is responsible for pneumonia, activating the immune response in the affected individual. In the lungs, inflammatory agents like neutrophils invade the alveolar spaces, causing lung cell clustering and reducing the efficiency of gas exchange. This inflammatory state may manifest as discomfort in the thoracic region, persistent coughing, elevated body temperature, and dyspnea. Bacterial agents such as Streptococcus pneumoniae, Haemophilus influenzae, and Staphylococcus aureus routinely emerge as significant factors in pneumonia incidents. The occurrence of viral pneumonia is often connected to influenza viruses, RSV, and adenoviruses. Pneumonia may present in a mild form, particularly when contracted from community sources, or in a severe form, as often observed in nosocomial settings. A chest X-ray allows for inspection of the lungs, heart, and blood vessels to determine the possibility of pneumonia infection. The chest X-ray for a healthy individual and an infected individual is shown in Fig. 1. Radiologists have major challenges when detecting pneumonia on chest X-rays due to the monochromatic color scheme. It is difficult to discern subtle changes in tissue density. Pneumonia can appear as lung field opacities in early or moderate cases, which can mimic normal structural structures such as the heart, ribs, and vascular systems. The visual signs of pneumonia may be obscured by overlapping anatomical structures and variations in image quality [7]. Unlike computed tomography scans, which provide detailed three-dimensional images, X-rays only provide a two-dimensional image, making it more difficult to distinguish pneumonia from other lung conditions. These factors highlight the potential effectiveness of deep imaging by contributing to the diagnostic challenges faced even by skilled radiologists. So, pneumonia detection through chest X-ray is a pending concern waiting to be resolved [8,9,10,11]. The technology-supported algorithms can aid human experts in reaching the diagnosis of pneumonia due to its reproducible decision support system and abundance of power.

**Fig. 1.**
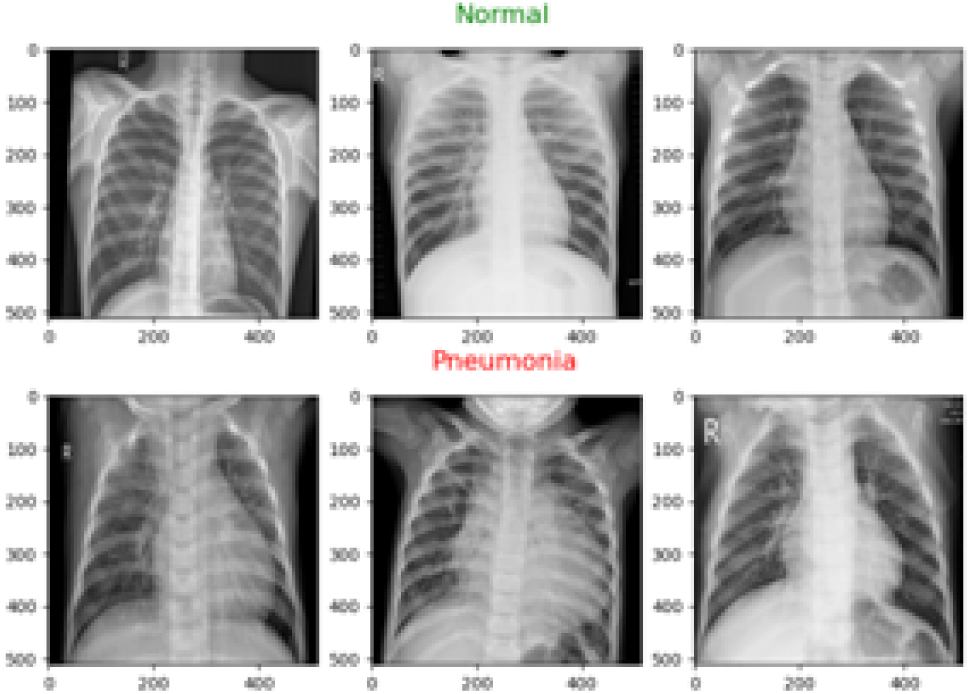
Chest X-ray scans of Healthy people vs Persons infected with pneumonia.

A meticulous diagnosis may be achieved through the utilization of precise imaging techniques and their subsequent interpretation. Established machine learning strategies can likewise serve for this goal [12,13,14,15]. These algorithms in machine learning execute modeling by utilizing features that have been meticulously extracted by specialists in the respective domain. A collection of machine learning techniques, including decision trees, random forests, K-nearest neighbors models, and an assortment of boosting methods, are engaged for detecting pneumonia through chest X-ray image analysis [12].

Traditional machine learning algorithms like Naive Bayes, Support Vector machines, and Random Forest run into obstacles in pneumonia discovery from X-ray scans fumbling with complex function design restricted capabilities for recording intricate spatial connections and vulnerability to heavy information concerns [16,17,18]. Their adaptability to progressive pneumonia situations as well as imbalanced datasets hinders their efficacy. On the other hand, CNNs can automatically learn hierarchical features and understand spatial relationships in images which has been proven to be highly effective in tasks like pneumonia detection using X-ray scans. Their capacity to handle raw pixel values, adapt to varied image sizes, and capture complex patterns makes them well-suited for medical image analysis [35,20,21,22].

Over the preceding ten years, deep learning has undergone substantial proliferation across a multitude of academic disciplines. Progress in fields such as materials science, cybersecurity, natural language processing, seismological studies, computational biology, and medical imaging analysis has been extraordinary [23,24,25,26,27,28]. Deep learning approaches, particularly convolutional neural networks, have utility in the recognition of pneumonia through the evaluation of chest X-ray radiographs. Through the utilization of extensive labeled datasets for training, CNNs acquire the capability to discern features that distinguish pneumonia from non-pneumonic cases. Pretrained architectures may undergo fine-tuning to enhance diagnostic precision.This methodology entails various stages of data preprocessing (featuring resizing, normalization, and augmentation), model training, and performance evaluation employing metrics such as accuracy, precision, and recall. Upon completion of the training phase, the model is primed for deployment within clinical settings, facilitating real-time diagnostic capabilities and assisting radiologists in the prompt identification of pneumonia.

This study delineates a Convolutional Neural Network (CNN) specifically designed for the detection of pneumonia in X-ray imaging, which incorporates semantic layers to facilitate the automated elimination of functions. Upon training with meticulously classified datasets, it demonstrates a high degree of accuracy in identifying pneumonia-positive cases and negative instances, employing metrics such as precision, accuracy, and recall as evaluative criteria. By concentrating on the optimization of hyperparameters and the application of sophisticated tuning methodologies, our CNN emerges as a dependable instrument for the effective diagnosis of pneumonia through radiographic images.

The remainder of the paper is structured as follows: Section 2 thoroughly examines contemporary studies on pneumonia detection for current insights. Section 3 offers a comprehensive exposition of the proposed methodology. Sections 4 and 5 emphasise the evaluation of the proposed solution and examine the documented findings. Section 6 summarises the outcomes of this study, whereas Section 7 outlines the prospective extensions of the current research.

## 2 Literature Survey

The comprehensive literature review pertaining to the research article on pneumonia detection meticulously analyzes contemporary methodologies, innovations, and challenges prevailing within the domain. The study conducted by Saul [29] implements a bifurcated approach to image processing preceding the modeling phase. This process significantly augments the visual attributes of images for the purpose of training. For classification, a convolutional neural network alongside a residual neural network is utilized. The research is predicated on a dataset comprising 4,000 X-ray images. The findings of this investigation assert a peak accuracy of 78.73% attained through modeling subsequent to the execution of preprocessing techniques involving contrast enhancement and illumination adjustments. Weighted classifiers are utilized within a methodological framework to categorize pneumonia based on digital representations of chest X-rays [30]. Utilizing advanced architectures in deep learning, we produce weighted predictions with models including InceptionV3, ResNet18, DenseNet121, Xception, and MobileNetV3. The dataset’s inherent bias is lessened by utilizing data enhancement methods. Their model achieves an F1 score and an AUC score exceeding 99. The researchers [31] developed an ensemble comprising two U-Net architectures, each leveraging distinct backbone networks—ResNet-34 and EfficientNet-B4. Their approach aims to amalgamate the strengths of both models to enhance segmentation performance. The U-Net design rooted in EfficientNet-B4 showcases remarkable efficacy in comparison to the ResNet-oriented U-Net design regarding pneumonia detection. The model encounters challenges in managing type-1 errors. The ensemble model attained a test accuracy of 90% on the evaluated dataset, indicating its potential effectiveness in clinical settings. The investigation acknowledges the imperative for further validation across diverse demographics and imaging conditions to ensure generalizability. Pneumonia can arise from either viral or bacterial pathogens. Rahman [32] investigates three classification categories using various pre-trained Convolutional Neural Networks. Transfer learning is employed to augment classification performance on a comparatively smaller dataset. The three classification categories encompass normal, pneumonia (both bacterial and viral), and pneumonia caused by viral and bacterial agents. The performance of DenseNet201 outshines that of earlier pretrained models, with accuracy figures hitting 98%, 93.3%, and 95% across the trio of classification systems, respectively. A smartphone application has been developed to assist in the detection of pneumonia [33]. CreateML is utilized for model development purposes. This application is capable of creating, training, and deploying models directly from a Mac. The study reports an accuracy of 85%.

The dataset comprising 100 X 100 images undergoes enhancement for traces through the application of the marker-controlled watershed segmentation methodology (MCW) [34]. These chest X-ray images are employed for the purposes of modeling. The process of modeling integrates attention mechanisms along with Long Short-Term Memory (LSTM) architectures. The methodology put forward sorts an image into one of three unique groups: COVID-19, normal, or pneumonia. For the purpose of pneumonia detection, the RSNA Pneumonia Detection Challenge dataset has been utilized. This dataset contains a total of 32,227 images [38]. The potential risk of overfitting is alleviated by the introduction of additional noise into the dataset. Furthermore, various enhancement techniques are employed. VGGNet variants with 19 and 16 layers, ResNet50—a residual neural network comprising 50 layers, InceptionNet v3, and YOLO v5 convolutional neural network models are utilized for machine learning applications. VGG16 attains the highest accuracy rate of 88%. The work [35] compared different deep learning models: a custom CNN, transfer learning with ResNet152V2, and a fine-tuned ResNet152V2. The fine-tuned ResNet152V2 model reported the test accuracy (96%), outperforming both the custom CNN (90%) and the standard transfer learning approach (85%). The study highlighted that false negatives (missed pneumonia cases) were more critical than false positives, emphasizing the need for high sensitivity. For feature extraction, the authors [36] used both the InceptionV3 model and a Deep CNN. To improve predictive accuracy, they used an entropy-normalized Neighborhood Component Analysis technique to reduce dimensionality, followed by an Ensemble-Modified Classifier using Naïve Bayes, XGBoost, and Random Forest algorithms. The ensemble model reported 92% accuracy.

Scholars worldwide are advancing the automation of pneumonia, COVID-19, and tuberculosis identification utilizing CT scans and X-ray imagery through the implementation of deep-learning frameworks [39]. This work [37] described Real-World Feature Transfer Learning (RWFTL), a method for embedding characteristics from natural images (ImageNet) for pneumonia detection. RWFTL optimizes feature adaptation from RGB photos to grayscale medical images by reproducing grayscale channels, allowing for seamless use of pretrained CNNs. This technique dramatically enhances model generalization, achieving 93.3% accuracy with InceptionNet-v3 and beating models trained from scratch. The work emphasizes that real-world features can bridge the domain gap between natural and medical images, providing a faster, more efficient deep learning strategy for medical diagnosis while minimizing reliance on large-scale annotated medical datasets. The highlights for a few prominent works are provided in Table 1. The following opportunities have been determined from the research gaps of the earlier works.

– The transfer learning approach with pre-trained models on medical imaging datasets, which could boost accuracy, is unexplored.
– With unbalanced data sets, the model may be biased toward the majority class. Discussion should be carried out on FP and FN cases.
– We need to check the computational efficiency and deployment challenges.

**Table 1.** The summarized details of prominent works in the area of pneumonia detection.

| Work | Dataset | Architecture | Results | Highlights |
| --- | --- | --- | --- | --- |
| [29] | Dataset of Radio-logical Society of North America: 4000 X-ray images | CNN RNN | Accuracy: 78.73% | Training set: 3000 images Testing set: 1000 images Pre-processing: increment in contrast and lightening methods |
| [30] | Large Dataset of Labeled Optical Coherence Tomography (OCT) and Chest X-Ray Images: 5836 X-ray images | InceptionV3, ResNet18, DenseNet121, Xception, and MobileNetV3 | Accuracy: 97.45%<br>Loss:0.087 | Data augmentation operations, Weighted classifiers |
| [31] | Kaggle dataset: 5863 X-ray images | Efficient Net-B4-based U-Net | Accuracy: 90%<br>Precision: 87%<br>Recall: 99% | Ensembled approach |
| [32] | Kaggle dataset: 5247 X-ray images of bacterial, viral, and normal chest X-rays | pre-trained CNN architectures: AlexNet, ResNet18, DenseNet201, and SqueezeNet | Accuracy: 98%<br>Precision: 97%<br>Recall: 99% | Data preprocessing, augmentation, and 3 classification schemes |
| [34] | ILD dataset: 1061 X-ray images | Attention-CNN LSTM | Accuracy: 96% | Preprocessing with MCW segmentation algorithm. |
| [35] | Kaggle dataset: 5863 X-ray images | Fine-tuned ResNet152V2 | Accuracy: 96% | The fine-tuned ResNet152V2 model outperformed the custom CNN and the non-fine-tuned transfer learning model. |
| [36] | Kaggle dataset: 5863 X-ray images | XceptionNet | Accuracy: 93% | Entropy-normalized Neighborhood Component Analysis (NCA) technique for dimensionality reduction. |
| [37] | Kaggle dataset: 5863 X-ray images | InceptionNet-v3 | Accuracy: 93.3% | Use of Real-World Feature Transfer Learning. |

We experimented with several architectures, including *VGG19*, *ResNet50*, *InceptionV3*, *DenseNet201*, and *MobileNetV3*, in order to fill the research gap in model effectiveness. A customized CNN was used as a benchmark for their performance.

Our CNN model was found to be the most effective in balancing accuracy, recall, and parameter complexity based on this comparative analysis. We used TensorFlow’s ImageDataGenerator for data augmentation in order to improve generalization and reduce dataset imbalance. Among the methods were shifting, zooming, and rotation. In order to ensure strong model performance across a range of inputs and enhance diagnostic consistency in practical settings, this increased the training set, enhanced class balance, and assisted in preventing overfitting. The trained model was locally deployed using TensorFlow’s Keras API, enabling real-time inference with a latency of less than 30 milliseconds per image. With minimal computational overhead and optimization for edge deployment, the system ensures quick, accurate predictions fit for clinical use without relying on expensive infrastructure or external servers.

## 3 Methods and Materials

### 3.1 Dataset

The current investigation employs Mendeley Data [40]. The dataset encompasses a total of 5,863 images. This compilation of information is structured into three unique directories: train, test, and validation, with every directory having two subfolders named Normal and Pneumonia, which contain X-ray images taken from a broad range of people. The training dataset consists of 5,216 images, systematically categorized into 1,341 X-rays from Normal patients and 3,875 X-rays from patients diagnosed with Pneumonia. Among the 624 images designated for testing, 234 are classified within the Normal category, while the remaining 390 are classified under the Pneumonia category. Located within the validation sub-directory, a collection of 16 images is present, consisting of 8 labeled as Normal and 8 labeled as Pneumonia. The dataset demonstrates a notable bias towards the Pneumonia category, necessitating the development of a strategy to rectify this imbalance. A viable approach to mitigate this bias involves the augmentation of the Normal class by incorporating additional image scans utilizing the Data Augmentation Technique [41,42,43].

### 3.2 Preprocessing and Augmentation

Data Augmentation constitutes a methodological approach that enhances the training dataset by producing modified iterations of the existing data. Image Augmentation represents a methodological technique that utilizes an image as the input and implements alterations that modify specific attributes of the original visual representation while maintaining its fundamental characteristics [44,45,46,47].

The augmentation function used in the paper is the ImageDataGenerator class from the TensorFlow. Keras.preprocessing.image module [48]. It has multiple hyperparameters in the method like featurewise_center, samplewise_center, featurewise std_normalization, samplewise_std_normalization, zca_whiteneing, zca_epsilon, rotation_range, width, height, brightness, shift range, horizontal and vertical flips, and many more. The preprocessing step involves utilizing hyperparameters like rescale, rotation range, zoom range, width, and height shift range to augment training data. The augmented images were reintegrated back into their respective sub-directories. The rotation parameter was set to 30 whereas the zoom range, width shift, and height shift ranges were set to 0.2, 0.1, and 0.1, respectively.

Subsequent to the preprocessing stage, specifically image augmentation as depicted in Fig. 2, the training subdirectory contained a total of 3875 images distributed across each subfolder. Each subfolder within the testing subdirectories was comprised of 390 images [11]. The validation dataset constitutes 20 percent of the training dataset, which was determined through a method of random sampling. Fig. 3 presents the balanced distribution of samples across classes in the dataset after the validation split.

**Fig. 2.**
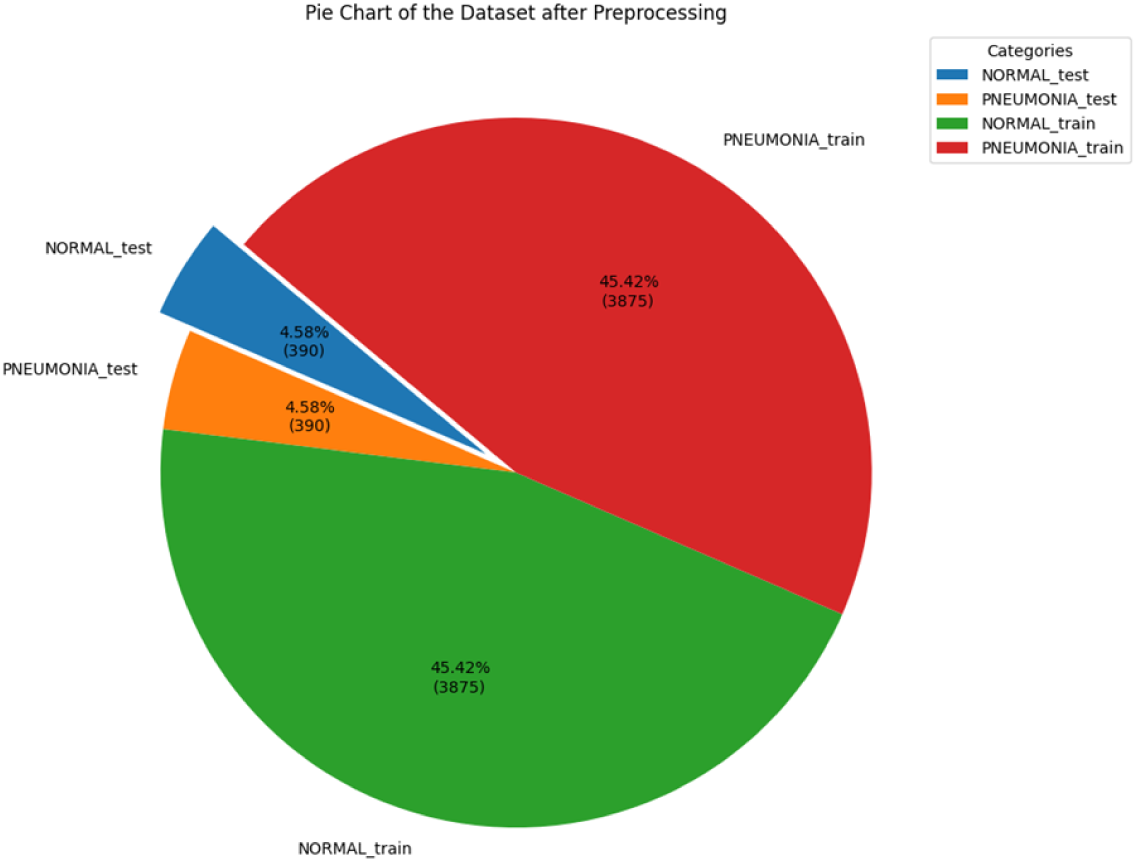
The pie chart delineates the balanced distribution subsequent to preprocessing but preceding validation split.

**Fig. 3.**
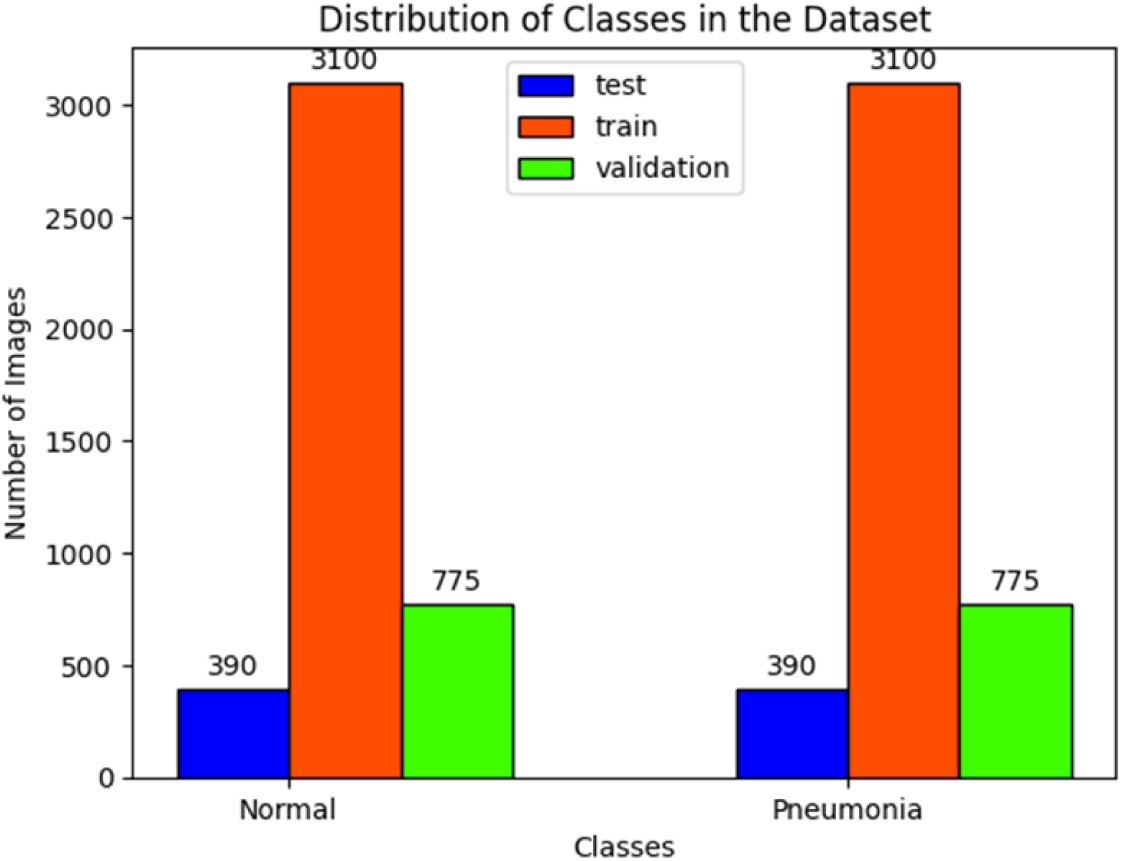
Bar Chart illustrating the post-validation split distribution of the balanced dataset.

### 3.3 CNN Architecture

The underlying architecture of Convolutional Neural Networks (CNNs) has appeared as a vital aspect in the landscape of deep learning, profoundly impacting domains including image classification, computational life sciences, language automation, and medical testing. CNNs are esteemed for their efficacy in image classification for several compelling reasons, as shown below.

– Dense Connectivity
– Enhanced Information flow
– Adaptability to different image sizes
– Parameter Sharing
– Feature Reuse and etc.

In the realm of CNNs, the filters used in convolution and the arrangement of layers are vital for capturing significant characteristics from visual content. The foundational layers are adept at identifying minute features, subsequently progressing to the more advanced layers, which discern larger features and intricate patterns [49,50,51,52]. This model encompasses 11 million parameters, which underscores its intricate nature and its capability to encapsulate nuanced details within image datasets. Figure 4 provides a comprehensive overview of the specifications pertaining to the customized Convolutional Neural Network (CNN) model. The color-coded three-dimensional representation of the bespoke CNN model depicted in Fig. 5 elucidates the convolutional block, max pooling layer, dropout layer, flattening layer, and dense layer. The convolutional layer at the beginning, referred to as the input layer, processes an input image that measures (256,256,3) and applies 32 filters with a kernel size of (3,3), using the ReLU for activation purposes. The subsequent convolutional block processes the outputs from the preceding layer, reducing the dimensions to 127×127 through the application of 64 filters with a (3,3) kernel, concurrently utilizing the ReLU activation function. Subsequently, a max pooling layer is introduced, utilizing a pooling size of (2,2) and accompanied by a dropout layer assigned a 0.2 dropout rate. The third, fourth, and fifth convolutional blocks proficiently manipulate the dimensions of input sizes measuring 62×62, 30×30, and 14×14, respectively. Each block manifests unique attributes with 128, 256, and 512 filters, all employing the ReLU activation function. In accordance with this framework, max pooling (2,2) and a dropout layer are systematically integrated subsequent to each convolutional block, thereby contributing to the classification of the network’s architecture. A dropout rate of 0.2 is consistently applied within the dropout layer. A new flattening layer has been added to ensure the outputs from the fifth convolutional block are standardized, along with a subsequent dropout layer set at a dropout rate of 0.2. Ultimately, a concluding dense layer comprising a single unit is incorporated, utilizing the Sigmoid activation function.

**Fig. 4.**
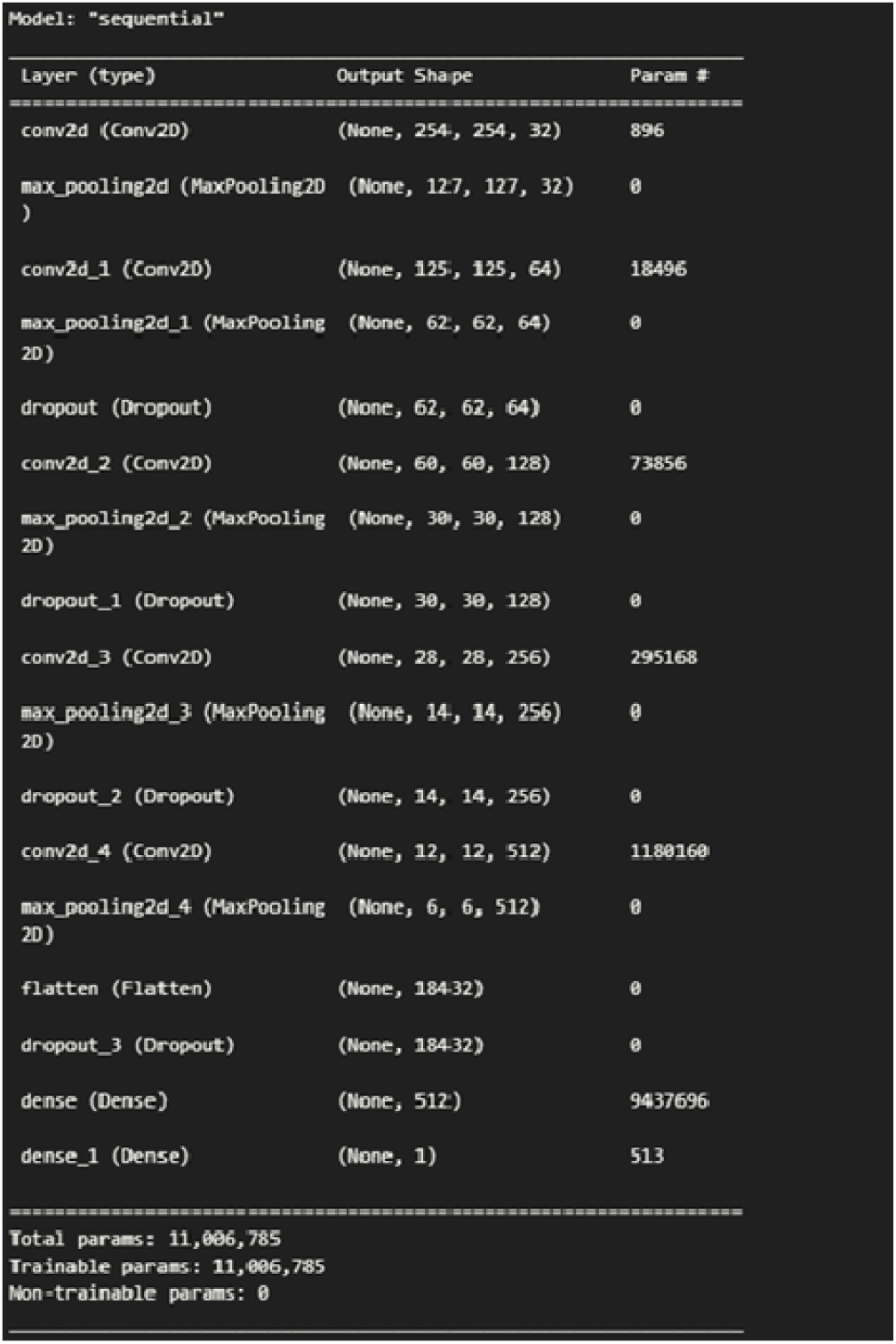
A concise summary of the architecture and characteristics of the custom Convolutional Neural Network (CNN) model.

**Fig. 5.**
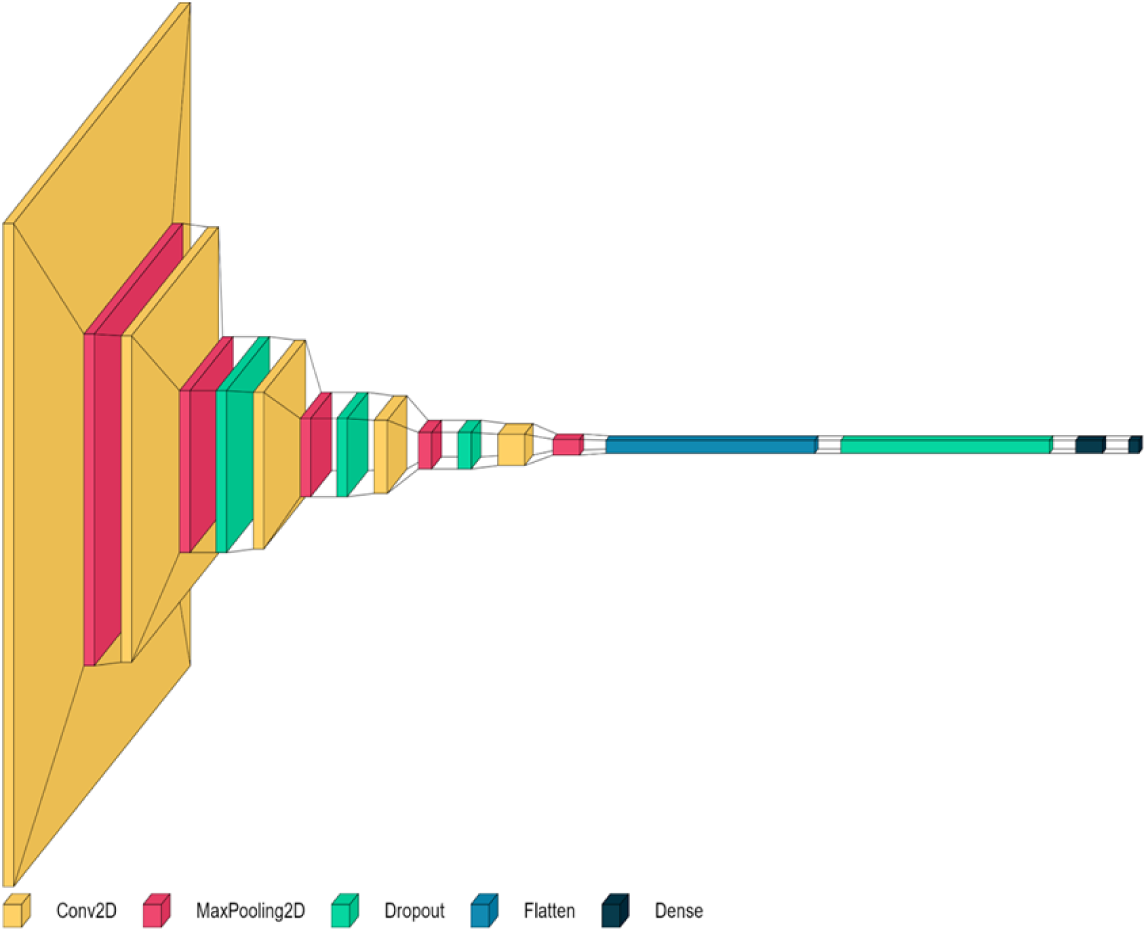
3D representation of the Convolutional Neural Network and the hidden layers of the custom model.

### 3.4 The Metrics

The metrics utilized for the validation of models are of equal significance to the other hyperparameters. These metrics elucidate the degree of accuracy achieved by the model. A wide selection of metrics is made available for the assessment of CNN models, featuring Accuracy, Precision, Sensitivity, False Positive Rate (FPR), and Recall, which are recognized as reliable signs. Accuracy assesses the overall correctness of the model and the accurate classification of instances. True Positive (TP) and True Negative (TN) demonstrate the accurately recognized positive and negative events. False Positive (FP) and False Negative (FN) reveal those that were mistakenly identified as positive or negative. When an instance is recognized as positive by the model and is factually a positive instance, it gets categorized as a True Positive (TP). In parallel, when a model pinpoints a negative instance correctly, it is marked as a True Negative (TN). Precision is ascertained by tallying the accurately identified instances and dividing this by the aggregate total of samples in the dataset, as represented in Equation 1. A case where the model mistakenly tags a true positive as negative is identified as a False Negative (FN) occurrence. Correspondingly, if a model inaccurately classifies a genuine negative instance as positive, it is designated as a False Positive (FP). The quantitative definition of precision is expressed as the fraction of true positive cases expected to the total count of positive predictions, as outlined in Equation 2. Recall, frequently called the true positive rate, is represented as the fraction of accurately identified positive examples to the complete tally of real positive examples, corresponding to Equation 3.

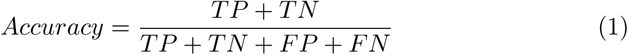

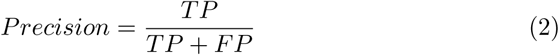

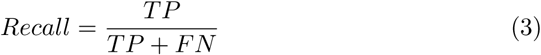

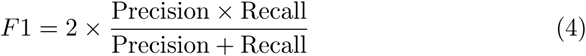

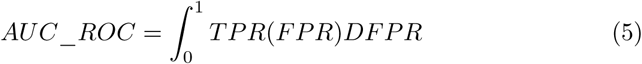

F1 score offers a balanced assessment of precision and recall, rendering it especially valuable in contexts where both measures are significant. The F1 score is determined by the harmonic mean of precision and recall as shown in Equation 4. The F1 score is very beneficial in binary classification tasks characterized by an uncommon class. The Area Under the Receiver Operating Characteristic Curve (AUC-ROC) is a commonly utilized statistic for assessing the efficacy of binary classification algorithms. It assesses a model’s capacity to differentiate between two classes at different threshold levels. AUC-ROC is a performance statistic that assesses the discriminative capability of binary classifiers by graphing the true positive rate versus the false positive rate across different threshold levels, as shown in Equation 5. The sensible factor to consider the metrics stands as an essential element in reviewing the efficiency of artificial intelligence designs. The F1 Score is the harmonic mean of precision and recall, balancing false positives and false negatives. It’s advantageous in imbalanced datasets where accuracy can be misleading. The AUC-ROC Curve illustrates the trade-off between true positive rate and false positive rate across thresholds. A higher AUC indicates better model discrimination between positive and negative classes.

These measurable procedures not only function as criteria for efficiency but also use important understandings that drive improvement methods that lift natural restrictions and help with constant formula improvement.

## 4 Training and Output

The visual representation of the process flow is shown in the Fig. 6. The model with the layers explained above was compiled with the optimizer as Adam with the learning rate set to 0.001. The Adam algorithm dynamically adjusts learning rates, addressing issues like vanishing or exploding gradients and expediting convergence. Also, the learning rate of the Adam optimizer was set to 0.001, and the loss function was Binary Cross Entropy. Accuracy, Precision, and Recall were taken as the metrics for the model.

**Fig. 6.**
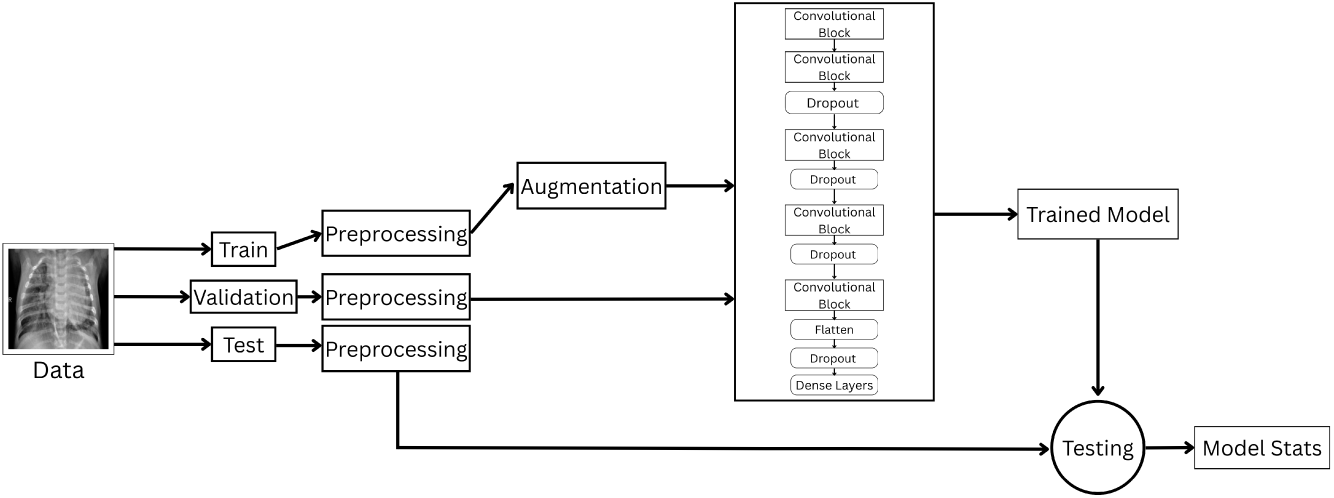
Visual representation of the process flow.

Fig. 7 showcases a graphical representation of the dynamic relationship between accuracy and the number of epochs for the Training and Validation phases. Fig. 8 depicts the progression of the Training and the Validation loss with the number of epochs. The model underwent a training regimen spanning 25 epochs, a temporal duration that was considered essential for attaining optimal performance with the balanced dataset, thereby resulting in exceptionally favorable outcomes. Upon the completion of 25 iterations, the training process culminated in a minimal training loss of 0.09, accompanied by a training accuracy of 97%. The evaluations for precision and recall indicated percentages of 98.3% and 97.64%, respectively. With a validation loss measuring 0.067, the validation accuracy reached 97.2 percent, and the precision and recall statistics were logged at 98.7 percent and 96.5 percent, respectively. The accuracy attained on the testing dataset, including 390 images for every class, was 96.5%, with precision measured at 94% and recall at 98%.

**Fig. 7.**
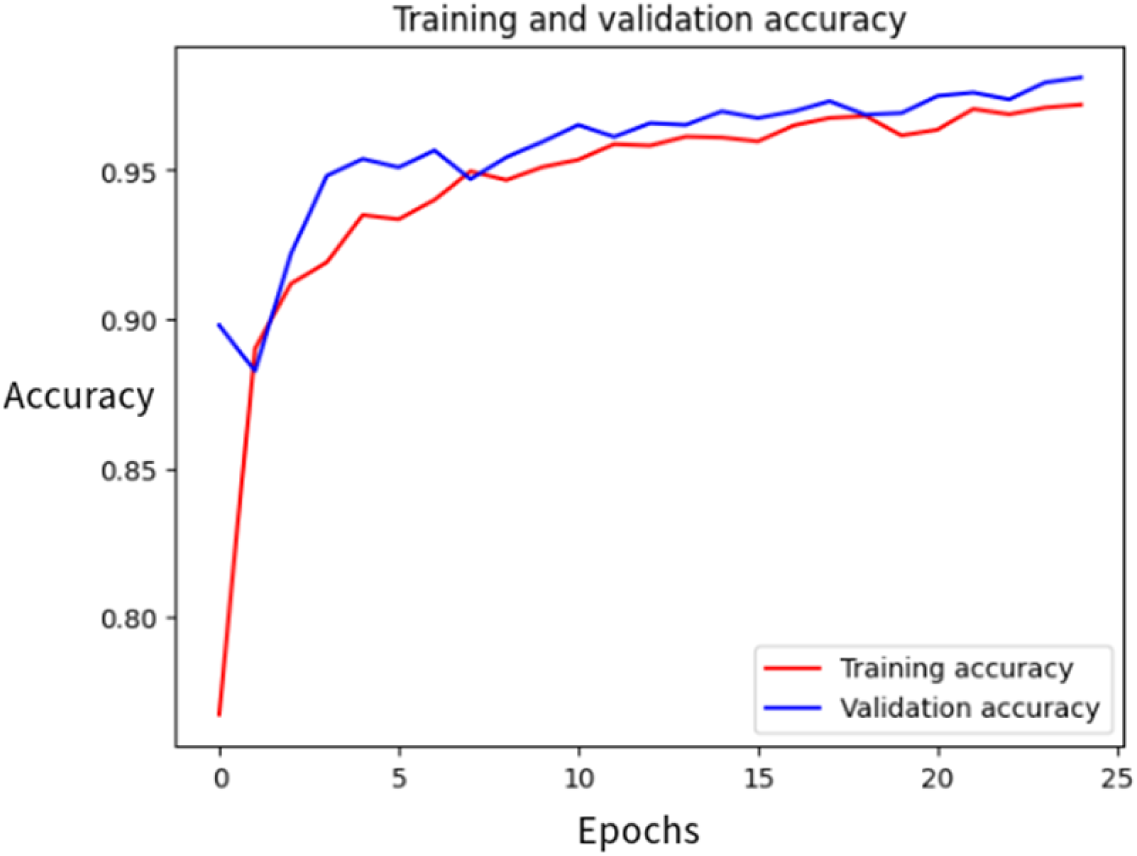
Graphical representation showcasing the dynamic relationship between accuracy and the number of epochs across both the Training and Validation phases.

**Fig. 8.**
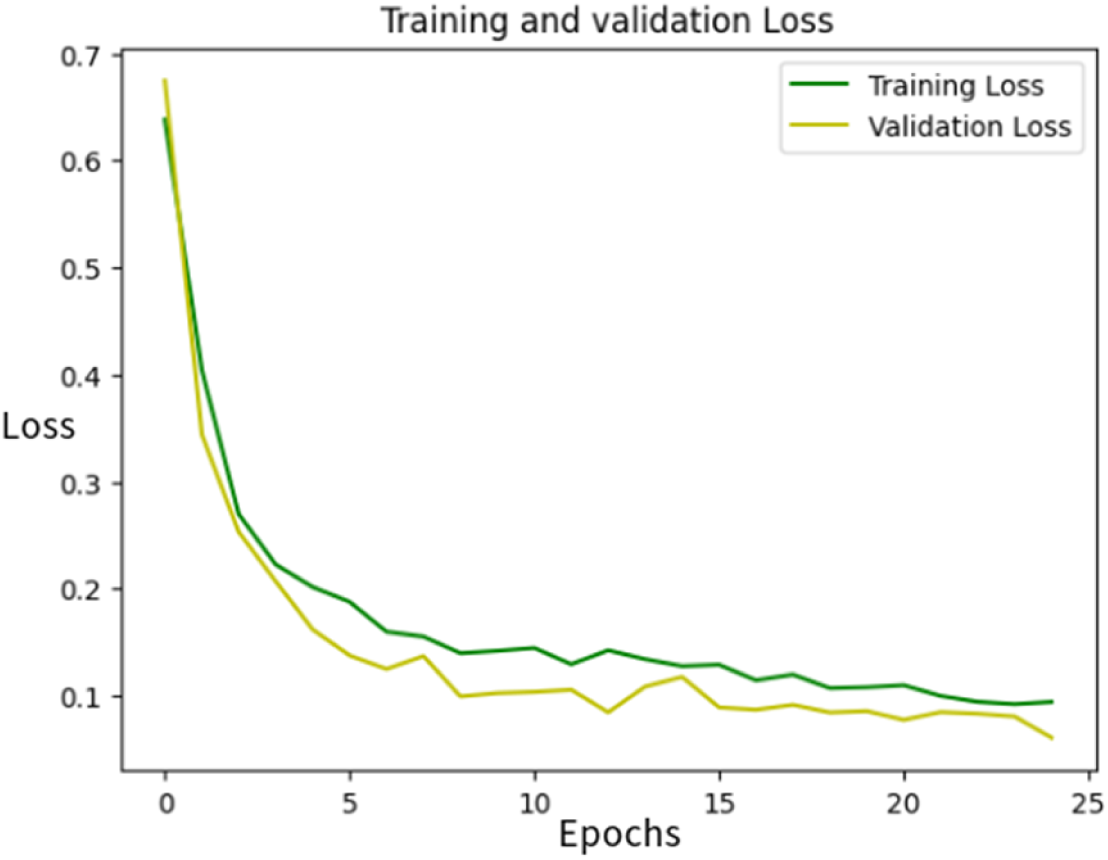
The progression of the Training and the Validation loss with the number of epochs.

Using an iterative learning strategy over more than 130 sampling iterations, the custom CNN model for pneumonia detection was trained with a strong focus on hyperparameter optimization. To determine the best configuration for the provided dataset, several crucial hyperparameters had to be systematically adjusted and assessed. The learning rate, batch size, dropout rate, number of filters per convolutional layer, and optimizer selection were the primary hyperparameters adjusted. A wide range of learning rates, with values ranging from 1 *×* 10*^−^*^5^ to 1 *×* 10*^−^*^2^, were investigated in early experiments. Given the binary classification nature of the task, a moderately low learning rate was chosen through this iterative refinement to guarantee stable convergence without overshooting the minima.

Another parameter that was thoroughly examined across a range of values, including 16, 32, and 64, was batch size. While larger batch sizes increased convergence speed but ran the risk of overfitting, smaller batches typically produced noisier gradients but frequently resulted in better generalization. In the end, a batch size that struck a balance between training stability and GPU memory efficiency was selected. Extensive research was also done on dropout rates, paying close attention to their distribution and size. A consistent value of 0.2 was found to offer the best trade-off between regularization and model capacity, successfully reducing overfitting without underutilizing the network’s representational power. Rates ranging from 0.1 to 0.5 were tested.

Iterations between Adam, RMSprop, and SGD were used to select the optimizer. Adam proved to be the most dependable, providing faster convergence and adaptive learning rates. In the early stages, the number of filters in each convolutional block was also thought to be a tunable parameter. A balanced architecture was required because, although increasing filter depth generally improved performance, it also had a significant impact on training time and memory usage.

To avoid overfitting and guarantee that performance gains were real and not coincidental, early stopping and validation monitoring were also included during hyperparameter searches. A combination of validation accuracy, loss, precision, and recall metrics was used to determine the final hyperparameter configuration, with recall being given priority because reducing false negatives is clinically significant. In addition to enhancing the model’s predictive capabilities, this meticulous, multi-round tuning procedure gave it a solid basis for reliable generalization in actual diagnostic situations.

After the threshold optimization of the classifier, i.e. the sigmoid function., with a threshold value of 0.68, the most optimized version of the model was achieved. With an accuracy of 96.5%, the precision and recall of 95% and 98.2% were achieved. Therefore, the confusion matrix appears as shown in Fig. 9

**Fig. 9.**
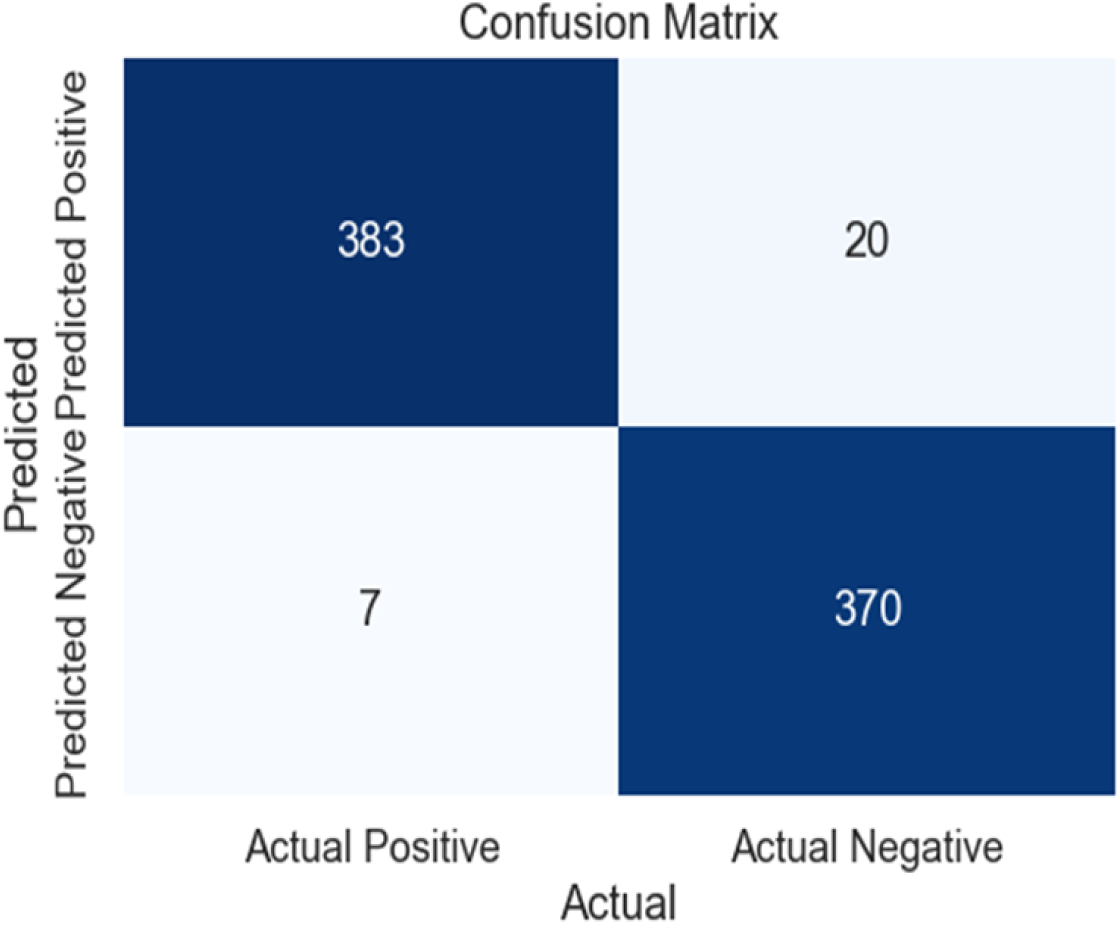
The confusion matrix delineating the conclusive results is displayed herewith.

The ROC curve, exhibiting an Area Under the Curve(AUC) of 0.98, portrays the classifier’s remarkable discriminatory efficacy as evident from Fig. 10. An area of 0.926 for the Precision-Recall (PR) curve signifies a superior performance of a binary classifier as shown in Fig. 11. It delineates the trade-off between precision (the accuracy of positive predictions) and recall (the capacity to identify all positive cases) at certain thresholds. A result of 0.926 indicates that the classifier is proficient in sustaining high precision and recall, which is very advantageous in the present context.

**Fig. 10.**
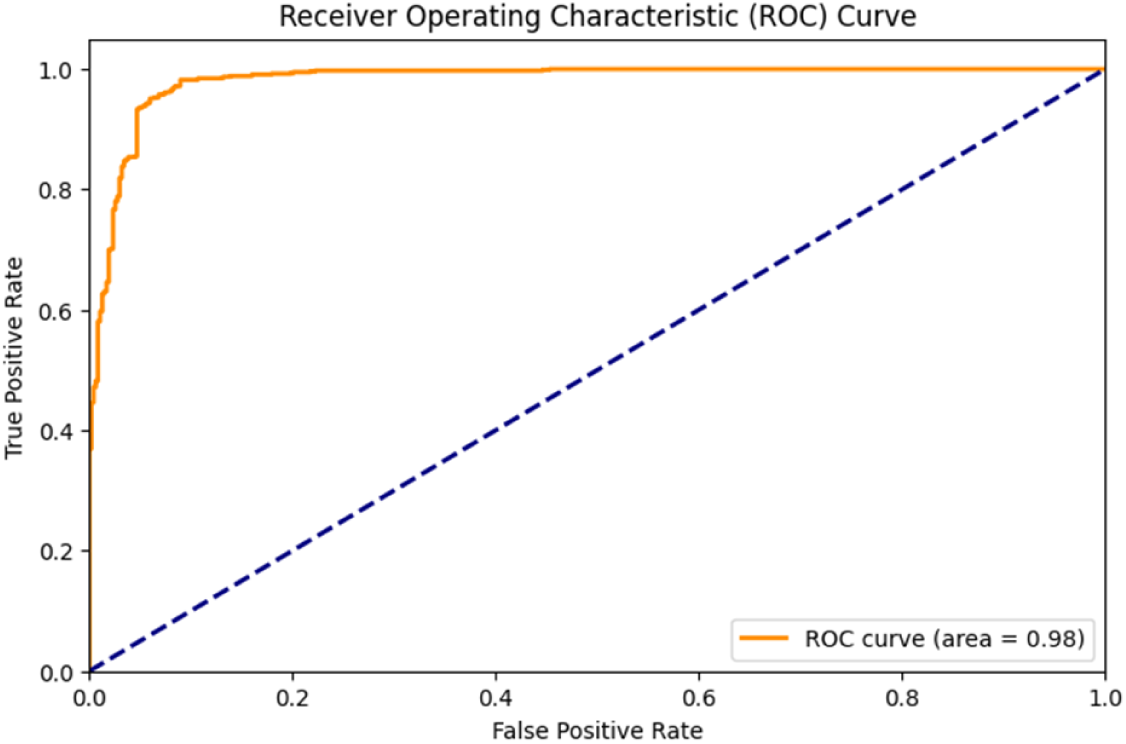
Visual depiction of the True Positive Rate (TPR) versus False Positive Rate(FPR) curve.

**Fig. 11.**
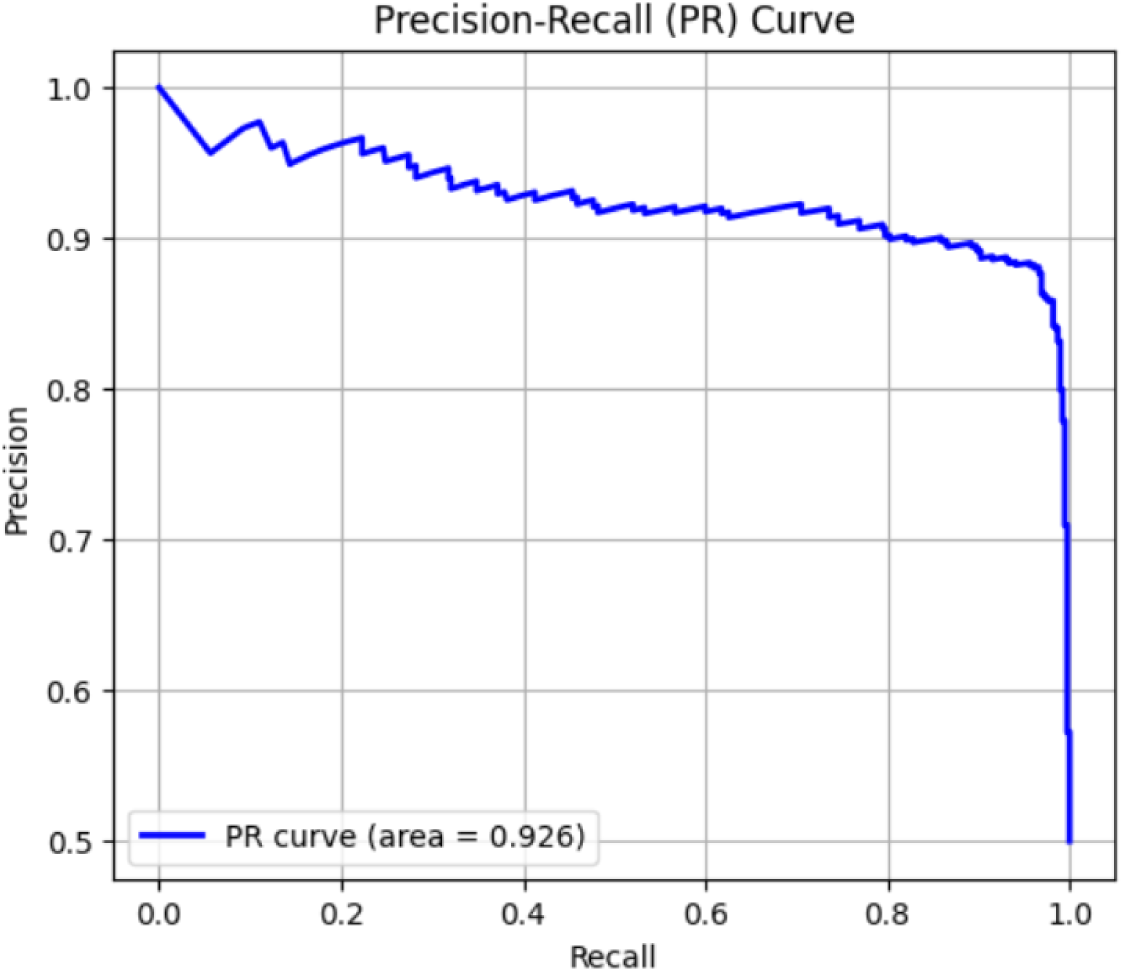
Visual depiction of the Precision Recall curve.

## 5 Discussion

This segment of the manuscript critically assesses the efficacy and scrutinizes the implications of all models that have been employed. The goal of this analysis is to systematically evaluate and rank five deep learning architectures. These architectures are VGG19, ResNet50, InceptionResNet50, InceptionV3, DenseNet201, and MobileNetV3. They are compared against a custom CNN utilizing a variety of metrics, inclusive of total parameters, training and validation losses, accuracy, precision, recall, and F1-score for training, validation, and test datasets, thereby yielding comparative insights between the proposed model and the pre-trained architectures. Figure 12 provides a graphical depiction of the models, showcasing accuracy across distinct phases, specifically Training, Validation, and Testing. Figure 13 presents the model losses over various time intervals. Figures 12 and 13 delineate the particulars of the leading three models: the proposed custom CNN, VGG19, and ResNet50. The quantitative synthesis of the various models trained through Transfer Learning is encapsulated in Table 2.

**Fig. 12.**
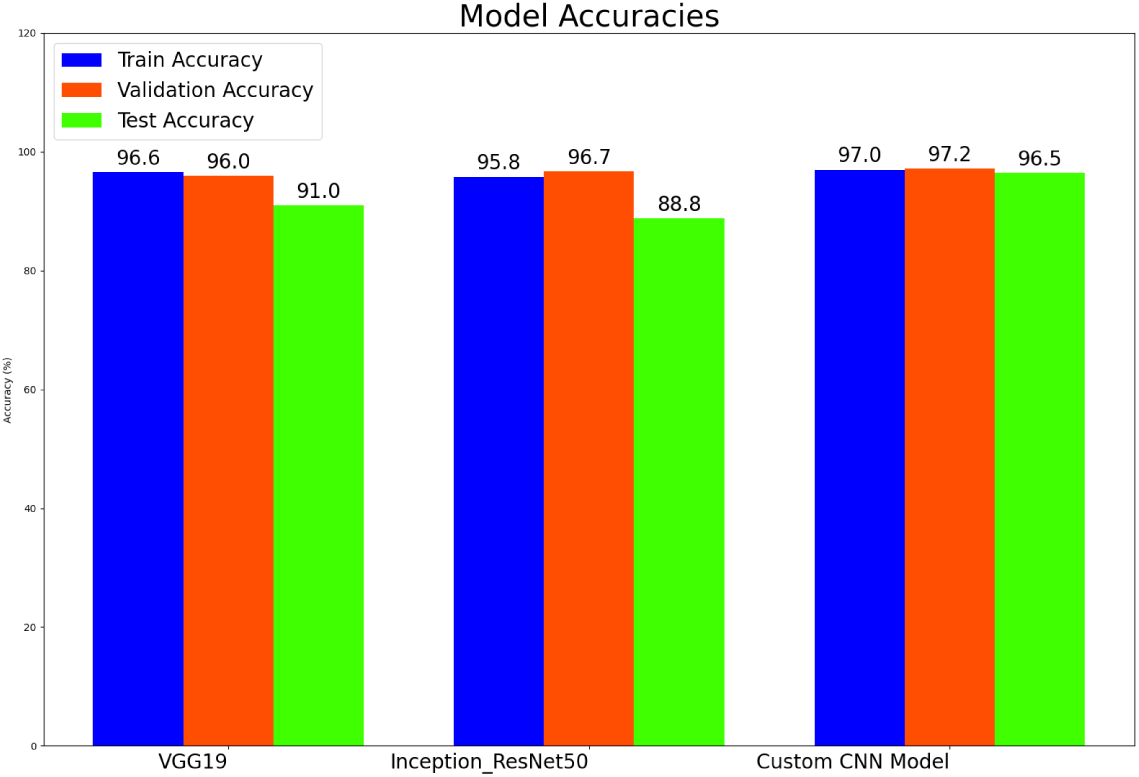
Graphic representation of accuracy for Top 3 models i.e. VGG19, ResNet50, and Custom CNN model during Train, Test, and Validate phase.

**Fig. 13.**
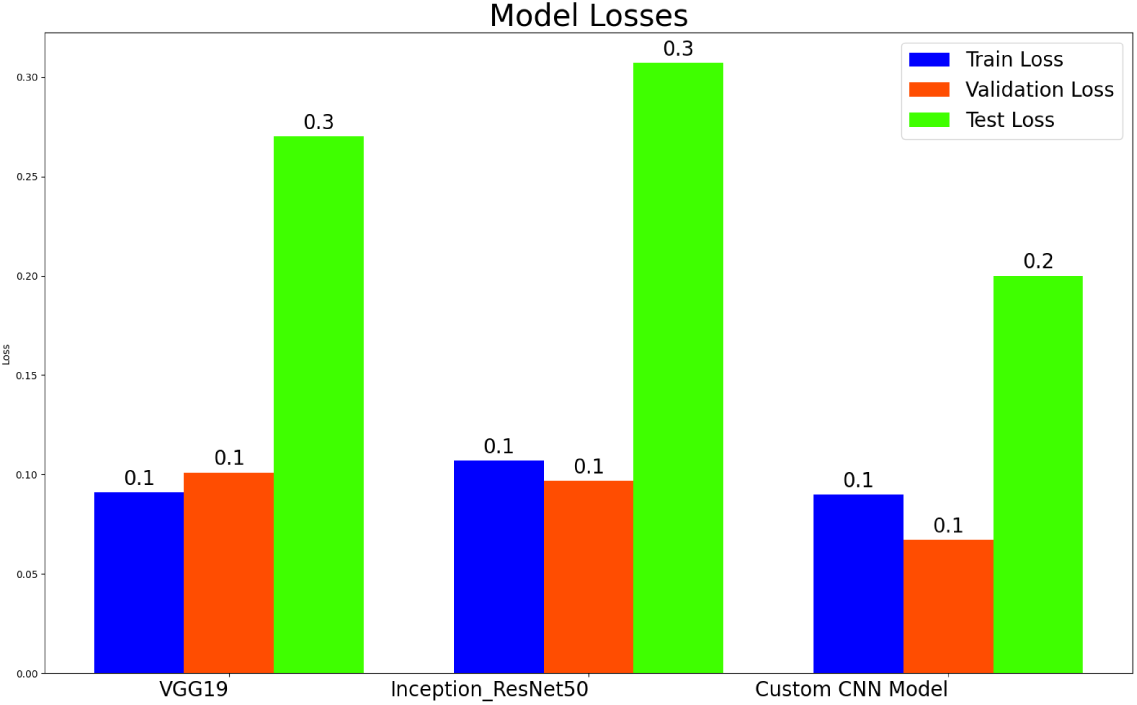
Visual depiction of Losses for Top 3 models i.e. VGG19, ResNet50, and Custom CNN model during Train, Test, and Validate phase.

**Table 2.** The summarized details of different models trained using Transfer Learning.

| Model | Total Params | Training Loss | Training Accuracy | Validation Loss | Validation Accuracy | Testing Loss | Testing Accuracy | Precision | Recall | F1 Score |
| --- | --- | --- | --- | --- | --- | --- | --- | --- | --- | --- |
| Custom CNN | 11M | 0.09 | 0.97 | 0.067 | 0.972 | 0.2 | 0.965 | 0.950 | 0.982 | 0.966 |
| VGG19 | 22.12M | 0.091 | 0.966 | 0.101 | 0.960 | 0.270 | 0.910 | 0.753 | 0.628 | 0.685 |
| ResNet50 | 31.97M | 0.281 | 0.897 | 0.222 | 0.918 | 0.339 | 0.867 | 0.250 | 0.500 | 0.333 |
| Inception ResNet50 | 57.87M | 0.107 | 0.958 | 0.097 | 0.967 | 0.307 | 0.888 | 0.5231 | 0.608 | 0.562 |
| Inception V3 | 26.50M | 0.118 | 0.956 | 0.134 | 0.946 | 0.348 | 0.865 | 0.550 | 0.501 | 0.524 |
| DenseNet201 | 26.19M | 0.066 | 0.977 | 0.046 | 0.985 | 0.633 | 0.816 | 0.669 | 0.582 | 0.622 |
| MobileNetV3 | 7.42M | 0.168 | 0.935 | 0.477 | 0.853 | 1.92 | 0.879 | 0.804 | 0.997 | 0.0.892 |

### 5.1 VGG19

VGG19 is characterized by a total of 22.12 million parameters. Throughout the training and validation phases, the model demonstrates a training accuracy of 96.6% alongside a validation accuracy of 96%. There exists potential for enhancement in the testing performance, with the current test accuracy being merely 91%. Nevertheless, with respect to the detailed metrics, VGG19 attains a precision of 75.3%, an F1-score of 68.5%, and recall rates of 62.8%, which suggests a slight omission of several favorable cases. The F1 scores across the validation, training, and testing datasets are disparate, which may signify a concern regarding overfitting.

### 5.2 ResNet50

The ResNet-50 architecture attained an accuracy of 89.7% during the training phase, accompanied by a loss metric of 0.281. The accuracy subsequently escalated to 91.8% during the validation stage; however, signs of overfitting became evident as the training accuracy surpassed that of the validation accuracy. Upon assessment with novel test datasets, it yielded an accuracy of 86.7% and a corresponding loss of 0.339. The model’s accuracy was quantified at 0.250. In contrast, its recall was determined to be 0.500, signifying that it successfully identified fifty percent of the actual positive instances. To furnish a holistic understanding of these metrics, we scrutinized the F1 score, which synthesizes precision and recall. The resultant score was 0.333.

### 5.3 InceptionResNet50

InceptionResNet50, with 57.87 million parameters, gets training and validation accuracies of 95.8%, and 96.7%, respectively, and a reasonable decline to 88.8% was recorded in testing accuracy. Its precision is 52.3%, recall is 60%, and F1-score is 56.2%, indicating moderate performance. A large number of parameters is not associated with better accuracy, which indicates the problem of decreasing returns to increasing model complexity.

### 5.4 InceptionV3

InceptionV3, comprising 26.5 million parameters, has training and validation accuracies equal to 95.6% and 94.6%, respectively. Its testing accuracy is 86.5%. InceptionV3’s precision is 55.0%, recall is 50.1%, and F1-score is 52. 4% which on the one hand suggests that it is a fairly good performing model, however, it is not the best of the rest, especially when precision and recall are major priorities in a given test.

### 5.5 DenseNet201

DenseNet201 [21], with 26M parameters, provides the highest training accuracy of 97.7% and validation accuracy of 98.5%. Nonetheless, on testing accuracy, it only achieved 81.7% and this was considered below average. Nevertheless, DenseNet201 defines relatively high precision, recall, and F1-score equal to 66.9%, 58.2%, and 62.2%, correspondingly. The results of testing where the model’s testing loss is a massive 0.634 show the generalization to unknown data, which needs some consideration with criteria; however, objective parameters show that DenseNet201 can be considered to be the best choice among the presented models, and further fine-tuning could be useful.

### 5.6 MobileNetV3

The effectiveness of the MobileNet model as a lightweight architecture was demonstrated by its 87.9% accuracy, 90% precision, and 99% recall for pneumonia detection. Pneumonia cases are rarely overlooked, thanks to the high recall, which is especially useful in medical diagnostics. The moderate accuracy and precision, however, suggest that although MobileNet provides speed, it might have trouble distinguishing pneumonia from other similar conditions. This illustrates how diagnostic performance and computational efficiency are traded off. Even though MobileNet is well-suited for real-time detection on mobile or edge devices, it may not be particularly well-suited for this use case.

The possibility of bias or level of generalization is also required to be checked. This assessment is carried out by training and evaluating model performance for different train, validation, and test data splits. It is carried out for different ratios, like 60-20-20, 80-10-10, and 75-15-10. The corresponding performance measure is reported in Table 3. This statistical test highlights two points: The model is generalized enough as the dispersion in the result is statistically less. This is due to preprocessing techniques applied to data and custom CNN architecture. This model can be used with other dataset provided same preprocessing applied on respective data to make it amenable to the requirement of custom CNN model.

**Table 3.** Performance measured for different dataset splits as part of statistical test.

| Split | Train Acc | Train Loss | Val Acc | Val Loss | Test Acc | Test Loss | Precision | Recall | F1-Score |
| --- | --- | --- | --- | --- | --- | --- | --- | --- | --- |
| 60-20-20 | 96.5 | 0.098 | 96.8 | 0.094 | 94.8 | 0.183 | 0.735 | 0.717 | 0.725888 |
| 80-10-10 | 96.3 | 0.108 | 96.5 | 0.1004 | 90.7 | 0.279 | 0.769 | 0.732 | 0.750044 |
| 75-15-10 | 96.3 | 0.1 | 97.15 | 0.081 | 87.2 | 0.43 | 0.786 | 0.742 | 0.763366 |

### 5.7 General Observations

The phenomenon of overtraining is discernible in all models that exhibit a deterioration in performance transitioning from the training/validation phase to the testing phase. Among the models analyzed, VGG19 demonstrates a relatively expedited processing time with a reduced number of parameters; therefore, the focus is placed more on the architectural design of the model rather than solely on the parameters themselves. As a result, it can be posited that none of the chosen metrics establishes a hierarchical advantage of one model over the others, suggesting that decisions regarding model selection should be predicated on the relative significance of diverse factors.

Accordingly, DenseNet201 has a good potential for fine-tuning and deployment because of its high training and validation accuracy and balanced accuracy, precision, recall, and f1-score. Despite this, the test loss hints at the model’s weakness in generalization, so it can benefit from regularization or data augmentation.

The findings suggest that the use of deep transfer learning, data augmentation, and ensemble modeling can significantly improve the accuracy of pneumonia detection in chest radiographs. Such models have the potential to serve as valuable tools in clinical settings, helping radiologists make quick and accurate diagnoses and ultimately contributing to better patient outcomes.

### 5.8 Model Deployment

After being saved in .h5 format, the trained CNN model was locally deployed using TensorFlow’s Keras API. The model was loaded without recompilation as part of the deployment pipeline, and the optimizer and metrics were reinitialized to match the training setup. To simulate batch input, input images were preprocessed by expanding dimensions, normalizing pixel values to the [0, 1] range, and resizing to 256*×*256 pixels. To perform real-time inference, Python’s time module was employed to measure latency. Suitability for low-latency applications was confirmed by a single prediction that averaged less than 25-30 milliseconds. To guarantee reproducibility, the deployment remained consistent with the training environment.

This configuration ensures data privacy and reduces runtime overhead—two important factors for medical applications—by enabling effective, local inference without the need for external servers or APIs. The pipeline is scalable and modular, allowing for integration into larger diagnostic systems. The whole project is available at here.

## 6 Conclusion

In conclusion, our examination of pneumonia detection leverages an in-depth understanding in conjunction with Convolutional Neural Networks and has manifested a considerable influence on clinical diagnostics. Employing a rigorously assembled dataset that encompasses reference sites for both normal and pneumonia instances, each additional component incorporated into the training analysis alongside recognition collections, characterized by an impressive 11 million parameters, has exhibited exceptional performance within this domain. Our model attains a distinctive screening accuracy of 96.5 percent, thereby establishing itself as a benchmark in the realm of pneumonia detection, exceeding the performance of existing bespoke model iterations. This accomplishment not only underscores significant advancements in clinical evaluation but also carries profound implications for the facilitation of early and accurate diagnosis. Our methodology effectively illustrates the capacity of CNNs to discern intricate patterns within radiographic images, while simultaneously affirming the resilience of our approach across varied datasets. In acknowledging this milestone, it is imperative to recognize the continuous evolution of contemporary healthcare technology. Future endeavors should concentrate on refining our design, identifying supplementary datasets, and addressing any inherent limitations. Our research exemplifies advancements in expert systems within the healthcare sector, underscoring the extraordinary potential of deep learning in the detection of pneumonia. This work contributes to academic discourse and holds promise for practical applications, bringing us closer to a future in which advanced technology revolutionizes clinical diagnostics.

## 7 Future Work

The following works can be carried out as an extension of the current work in medium to long durations:

– The model’s performance on X-ray images from different hospitals, geographic regions, and diverse populations can be tested.
– Use cross-validation on multiple datasets and assess the model’s performance on unseen hospital data.
– Extend the model to multi-class classification to distinguish between different types of pneumonia.
– Implement adversarial training techniques to make the model robust to real-world noise and perturbations.
– Optimize models for edge computing and mobile deployment to enable usage in low-resource settings.
– Implement XAI techniques to provide better insights into model decision-making.

## Data Availability

All data produced are available online at Kaggle

https://github.com/MalayVyas/PneumoniaApplication

## Author contribution

Malay Vyas: Conceptualization, Investigation, Writing. Apurva A. Mehta: Visualization, Writing, Supervision.

## Data Availability Statement

Not Applicable

## Research Involving Human and/or Animals

Not Applicable

## Informed Consent

Not Applicable

## A Traditional Machine Learning and Deep Learning

The juxtaposition of classical machine learning (ML) and deep learning (DL) elucidates notable disparities in their techniques, applications, and performance attributes. Conventional machine learning models generally depend on feature extraction and less complex methods, rendering them efficient for structured data contexts. Conversely, deep learning models, defined by their multi-layered neural networks, demonstrate superior performance in managing extensive, unstructured datasets, particularly in picture and speech recognition applications. This differentiation is essential for choosing the suitable method according to data type, computing capabilities, and particular application requirements.

### A.1 Methodology and Complexity

Traditional Machine Learning includes methods like Support Vector Machines, Random Forests, and K-Nearest Neighbors, which require manual feature extraction and selection. These approaches are generally more direct and enhance interpretability, making them suitable for structured datasets and situations where understanding is crucial.

Deep Learning utilizes neural networks with several layers (e.g., Convolutional Neural Networks, Recurrent Neural Networks) that independently extract features from raw input. This functionality allows Deep Learning models to adeptly handle complicated patterns and unstructured datasets; nevertheless, it simultaneously increases the complexity and computational requirements of the models.

### A.2 Performance and Applications

Standard Machine Learning shows enhanced effectiveness in activities that employ structured data, such as classification, regression, and clustering. It often provides improved consistency in specificity and overall accuracy, as evidenced by its applications across diverse domains.

Deep learning frameworks illustrate notable competence in assignments that demand strict precision and recall, particularly in realms like picture and sound recognition, language processing, and imaging analysis in medicine. Deep learning architectures have shown considerable effectiveness in detecting cognitive impairments and facial recognition, often exceeding the capabilities of human experts in specific areas.

### A.3 Generalization and Extrapolation

Traditional Machine Learning generally exhibits adequate performance within the parameters of the training dataset; nevertheless, it may struggle to extrapolate data outside the defined training area. This limitation might profoundly affect the model’s relevance in practical situations when the data may be incomplete or exceed the observed ranges.

Deep Learning demonstrates inherent skills that enable generalization beyond the training environment, making it more robust for applications requiring extrapolation. This attribute is essential for tasks where the model must infer data points beyond the training domain’s parameters.

### A.4 Challenges and Considerations

In terms of interpretability and computing efficiency, conventional machine learning has notable benefits. Its use with unstructured data, however, sometimes calls for considerable feature engineering and may show poor performance.

Deep learning Though they may learn characteristics on their own, deep learning systems demand significant computer power and vast data sets. The problems connected to these models include model selection, interpretability, and long training times.

To sum up, while deep learning offers great advantages in handling complex, unstructured data and achieving high accuracy, traditional machine learning remains a relevant approach for structured data environments and scenarios where interpretability and computational efficiency are crucial. The particular needs of the work, data characteristics, and available resources should guide the use of these methods. Furthermore, hybrid models combining both approaches are developing to use the benefits of each to address several challenges in artificial intelligence.

